# Plasma proteome-based test (PROphetNSCLC) predicts response to immune checkpoint inhibitors (ICI) independent of tumor programmed death-ligand 1(PD-L1) expression and tumor mutational burden (TMB)

**DOI:** 10.1101/2024.09.09.24313374

**Authors:** Yehuda Brody, Ben Yellin, Itamar Sela, Yehonatan Elon, Igor Puzanov, Hovav Nechushtan, Alona Zerkuch, Maya Gottfried, Rivka Katzenelson, Mor Moskovitz, Adva Levy-Barda, Michal Lotem, Raya Leibowitz, Yanyan Lou, Adam P. Dicker, David R. Gandara, Kimberly McGregor

## Abstract

Despite the approval of PD-(L)1 inhibitors for the first-line treatment of all metastatic, driver-negative, non-small cell lung cancer patients (mNSCLC) in the United States since 2018, there still is a lack discerning biomarkers to predict which patients will derive significant benefit. Tumor expression of programmed-death ligand 1 (PD-L1), measured as the tumor proportion score (TPS), is a standard biomarker approved for the selection of initial therapy. Tumor mutational burden (TMB), a promising biomarker, thought to represent the tumors’ ability to engage the host’s immune system, has demonstrated clinical utility primarily in the context of immunotherapy monotherapy. PROphetNSCLC, a test developed through proteomic analysis and machine learning, provides a novel approach by capturing biological processes from both tumor and host. In a previously published study, PROphetNSCLC, was validated to correlate with the probability of clinical benefit, independent of but also complementary to PD-L1 expression levels predicting specific treatment-related survival outcomes. Utilizing available tumor TMB measurements from this investigation, we sought to assess the correlation between the PROphetNSCLC clinical benefit probability score and TMB measurement. PROphetNSCLC demonstrated a correlation with various outcomes from PD-(L)1 inhibitor treatment independent of TMB status, whereas TMB did not exhibit an association with outcomes. This finding emphasizes the significance in of novel systemic biomarkers in refining personalized treatment strategies for mNSCLC.

## Introduction

The advent of immune checkpoint inhibitors (ICIs) has significantly altered the treatment landscape for metastatic non-small cell lung cancer (mNSCLC), providing a novel therapeutic approach for patients with historically poor prognoses. However, despite the considerable potential of ICIs, their efficacy varies substantially among patients, with only a minority having durable responses. This variability emphasizes the critical need for reliable biomarkers to predict which patients are most likely to benefit from ICI therapy. Tumor measured Programmed Death-Ligand 1 (PD-L1) proportion score (TPS) currently remains the sole recommended biomarker for ICI use in mNSCLC (NCCN Clinical Practice Guidelines in Oncology (NCCN Guidelines®) Non-Small Cell Lung Cancer, Version 7.2024). However, even with the harmonization and validation of the immunohistochemistry (IHC) assay for PD-L1 it still lacks sensitivity and specificity, necessitating the investigation of alternative or complementary biomarkers (Merino et al., 2020). Tumor mutational burden, the measurement of somatic mutations as detected by next-generation sequencing, is hypothesized to be a surrogate measure of potential neoantigens presented to the host to elicit an immune response and thus correlate with responsiveness to ICIs. This hypothesis was evaluated and corroborated through multiple trials across various tumor types (Sholl et al., 2020) and subsequent studies demonstrated that PD-L1 and TMB were distinct biomarkers (Carbone et al., 2017; Hellmann, Callahan, et al., 2018). TMB was then recognized as an emerging biomarker for mNSCLC patients based on the initial findings of prolonged progression-free survival in the CheckMate 227 study. However further updates from this trial later revealed improved outcomes from ICI treatment irrespective of TMB, PD-L1 or a combination of both (Hellmann, Ciuleanu, et al., 2018; Hellmann et al., 2019). This study along with several other conflicting studies along with concerns related to the technical components of the biomarker (Carbone et al., 2017; Rizvi et al., 2018) provided evidence for the removal of TMB as an emerging biomarker from the NCCN guidelines in 2020 (Ettinger et al., 2021). Later studies also noted concerns about the reliability of PD-L1 despite it being a more technically feasible biomarker (Ge et al., 2022; Khagi et al., 2017; Pérol et al., 2022; Zhou et al., 2019). This was particularly noteworthy given the absence of randomized controlled trial data to further guide regimen selection between PD-(L)1 inhibitor monotherapy and PD-(L)1 inhibitors combined with chemotherapy in this population, and real-world meta-analyses have yielded conflicting results (Ge et al., 2022). PROphetNSCLC, a novel proteome-based biomarker test, has been clinically validated to predict clinical benefit to PD-(L)1 inhibitors in mNSCLC patients independently of tumor PD-L1 expression levels and was associated with significantly different outcomes for patients with PD-L1 high tumors treated with monotherapy versus combination therapy (Christopoulos et al., 2024).

Despite the challenges of TMB as a biomarker for initial treatment selection for mNSCLC patients, TMB is routinely reported as part of comprehensive genomic profiling and continues to be of interest and included in ongoing research (Galvano et al., 2021; Merino et al., 2020; Ricciuti et al., 2022). Some of the limitations around TMB were related to the tissue requirements, various methodologies, lack of standardization, and complex reporting, which were also seen in the early utilization of PD-L1 staining by IHC (Sholl et al., 2020). Notwithstanding the improvements towards standardization of these processes, both biomarkers still exhibit notable limitations for predicting response to immunotherapy, with TMB demonstrating a lack of clear data for treatment guidance no in the first-line setting and no utility when considering any ICI combination regimen. PROphetNSCLC, presents a significant advancement, by utilizing a blood-based test to capture both tumor and host components and employing machine learning to analyze the proteomic profile of an individual patient, in order to predict their probability of clinical benefit from PD-(L)1 inhibitor-based therapy alone or in combination with chemotherapy. This score, when combined with PD-L1 as a composite biomarker, can inform treatment decisions in the first-line setting where multiple options exist. In our large prospective observational trial, PROPHETIC (ClinicalTrials.gov identifier: NCT04056247), which provided the patient population for the development and validation of PROphetNSCLC, we aimed to explore the correlation between TMB and PROphetNSCLC and clinical outcomes in our real-world ICI treated patients and compared them with PROphetNSCLC results.

## Methods

### Study Design and Patient Selection

From patients enrolled in a multicenter observational study PROPHETIC (ClinicalTrials.gov identifier: NCT04056247), across 20 centers in Israel, Germany, the United Kingdom, and the United States between January 2016 and July 2022, a cohort of 472 driver-negative mNSCLC patients treated in the first line with either a PD-(L)1 inhibitor or a PD-(L)1 inhibitor in combination with chemotherapy were identified and reviewed for inclusion of next-generation sequencing assessed tumor mutational burden (TMB) reported numerically from (muts/Mb), TMB High (>10 muts/Mb) or Low (<10 muts/Mb), and whom also had a pretreatment blood sample available for PROphetNSCLC testing as previously described (Christopoulous, et al 2024). Plasma and data collection were also previously described and briefly included using double-spun plasma, electronic medical record extraction with source validation, physician’s choice PD-L1 testing, and documented treatment and physician response assessment. Proteomic measurements were obtained from plasma using the SomaScan Assay v4.1, which uses Slow Off-Rate Modified Aptamers, and analyzed using the validated and locked down machine learning-based algorithm (Yellin et al., 2024).

### Definition of Clinical Benefit, PROphetNSCLC, PD-L1 and TMB

Clinical benefit (CB) was defined as no evidence of progressive disease (PD) within 12 months after treatment initiation based on radiographic imaging according to RECIST version 1.1, or other clinical criteria consistent with PD based on physician assessment. The PROphet model assigned a score of 0-10, which correlated with a probability of CB, and this was converted into a binary result of ‘positive’ or ‘negative’ based on threshold set during development of 0.27 for reporting of the PROphetNSCLC CB probability prediction. Tumor mutational burden (TMB) was categorized as “High” if >10 muts/Mb or ‘Low’ if < 10 muts/Mb as reported by the site treating physicians (Ready et al., 2019). PD-L1 scoring was classified as negative (<1%), low (1-49%), or high (>50%).

## Results

65 mNSCLC patients treated with a PD-(L)1 based regimen with both TMB and PROphetNSCLC results available, were similar to the total cohort of 444 mNSCLC patients treated in the first line setting with a PD-L1 inhibitor alone or in combination with chemotherapy (Supplemental Table 1, Supplemental Table S2-patient data). Of these, patients treated with a PD-(L)1 inhibitor containing regimen who were PROphetNSCLC POSITIVE had a significantly improved real world overall survival (rwOS) compared to those who were PROphetNSCLC NEGATIVE (rwOS 23.8 mo *v* 10.2 mo; HR, 0.36 [95% CI:0.18-0.71]; p= 0.0033; Fig 1A) while on the contrary TMB status (TMB-High >10 muts/Mb or TMB-Low <10 muts/Mb) was not statistically significantly correlated with rwOS (rwOS 17.1 m *v* 12.8 mo; HR, 0.99 [95% CI: 0.52-1.88];p=0.9837; Fig 1B). When looking at correlation of TMB between those treated with a PD-(L)1 inhibitor monotherapy (n=30) or when combined with chemotherapy (n=35)(Supplemental Fig 1A,B) there was still no significant association, however PROphetNSCLC POSITIVE patients had a significant rwOS when treated with PD-(L)1 inhibitor monotherapy compared to PROphetNSCLC NEGATIVE patients (rwOS 23.8 m v 11.1 mo; HR, 0.30 [95% CI: 0.12-0.78];p=0.0127; Supplemental Fig 1C,D). Additionally, in all evaluable patients who had a tumor where TMB was reported we found no correlation between TMB High or Low or numerical value with the PROphetNSCLC CB Prediction Score (Fig 1C,D). A univariate analysis showed that PROphetNSCLC and PD-L1 expression are predictive yet independent, while TMB is not (Figure 2).

**Figure 1:**
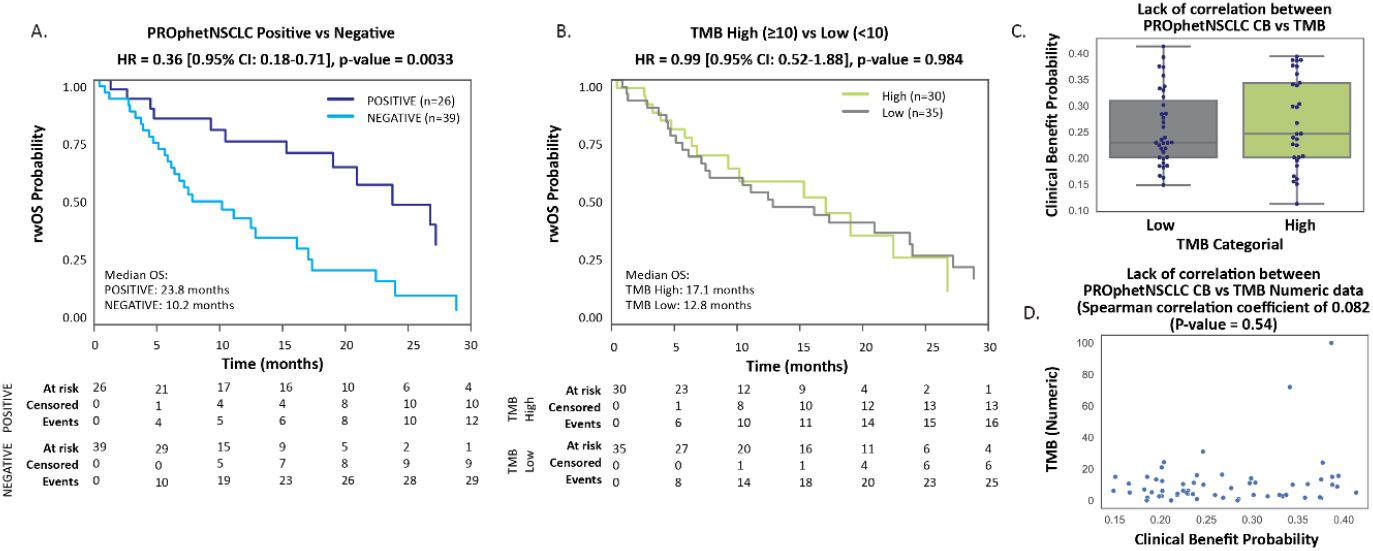
A. Kaplan-Meier plot, PROphetNSCLC result correlation with differential rwOS outcomes B. Kaplan-Meier plot demonstrating that TMB alone does not correlate with rwOS outcomes. Plot of PROphetNSCLC clinical benefit (CB) probability score separated by TMB groups C. Boxplot and swarm (TMB High [>10 muts/Mb], Low [<10muts/Mb]): each point representing a patient D. Scatter plot on the TMB numeric value demonstrating the independence of PROphetNSCLC CB probability and TMB in continuous values.

**Figure 2:**
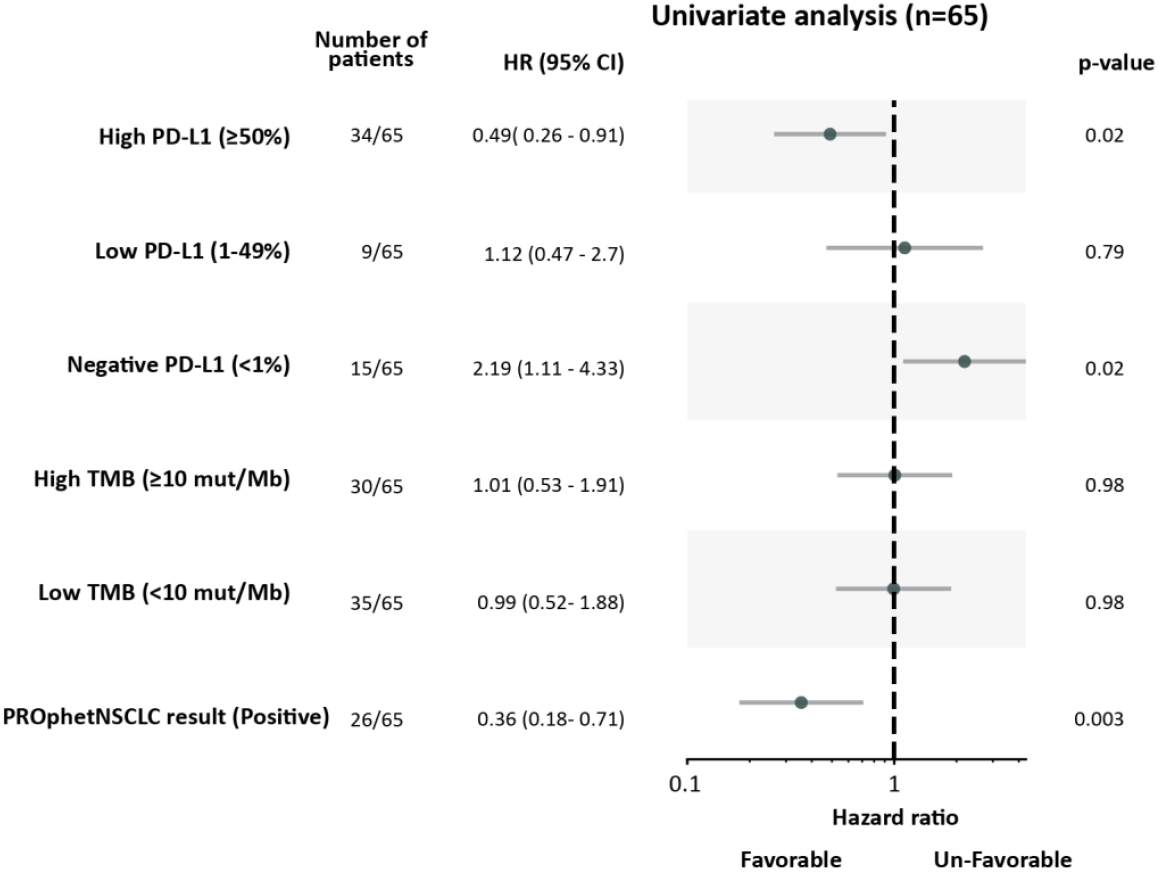
Univariate Analysis on the TMB cohort set (n=65) Forest plot illustrating the univariate analysis on the discussed biomarkers PD-L1 TMB and PROphetNSCLC. Each row represents a variable, with lines indicating the 95% confidence intervals (CIs) and corresponding hazard ratios (HRs). The central vertical dotted line signifies the HR for the null hypothesis (HR = 1), acting as a reference point for assessing the impact of each variable on survival.

## Discussion

The unmet need for improved biomarkers for immune checkpoint inhibitors (ICI) responsiveness in metastatic non-small cell lung (mNSCLC) cancer is well described. Tumor mutational burden (TMB), hypothesized to correlate with tumor immunogenicity which could be transformed into an anti-tumoral host immune response, has inadequate data and technical challenges preventing it from currently being recommended in the NCCN guidelines as a biomarker for the first-line treatment considerations in mNSCLC. Even when evaluating TMB performance in immunotherapy-only regimens for first-line mNSCLC, such as Checkmate 227, further follow-up demonstrated no correlation with outcomes from either TMB, programmed death-ligand 1 (PD-L1) levels, or a combination of the two biomarkers.

TMB has also consistently been shown to have no predictive value for the more commonly used PD-(L)1 inhibitor + chemotherapy combinations for either squamous or non-squamous mNSCLC patients(Paz-Ares et al., 2019; Sesma et al., 2020). Despite ongoing research in better characterization of mutations contributing to TMB and the potential for host engagement, TMB categorized as ‘High’ and ‘Low’ using a threshold of 10 muts/Mb, has no role as a standalone biomarker for first line mNSCLC treatment selection. Nonetheless, given the more widespread utilization of comprehensive genomic profiling and reporting of TMB measurement and supporting data, there remains ongoing consideration by physicians in practice. Moreover, despite broad adoption of tumor testing for PD-L1 expression, and studies indicating patients with PD-(L)1 high tumors may be spared the additional toxicity of chemotherapy added to PD-L1 inhibitors, this practice has not been widely adopted by practicing physicians. This hesitation is attributed to the absence of a randomized clinical trial comparing PD-(L)1 monotherapy and combination, conflicting evidence across three large real world meta-analysis, and a more recent FDA pooling of several randomized controlled trials (Goulart et al., 2024) which could create concerns in the utilization of PD-L1 expression alone as a biomarker.

PROphetNSCLC, a test that captures biological responses of both host and tumor by measuring levels of functional proteins, has the potential to provide novel insights not attainable from tumor-derived PD-L1 expression and simplified TMB measurements. This is substantiated by our studies demonstrating that PROphetNSCLC is both independent of PD-L1 expression levels and TMB. Our results were not unexpected, given TMB has not been shown to have predictive value in chemoimmunotherapy-treated patients; furthermore, we observed no correlation between TMB and survival outcomes in patients treated with PD-(L)1 inhibitors alone. PROphetNSCLC, conversely correlated with overall survival independently of TMB and was able to differentiate outcomes based on either a PD-(L)1 inhibitor alone or combined with chemotherapy in patients with tumors exhibiting high PD-L1 >50% expression. Additionally, it predicted statistically significant survival outcomes when treated with PD-(L)1 + chemotherapy combination regimens in those with a PD-L1 tumor expression level of <50%. In conclusion, the PROphetNSCLC test provides individualized insights from both tumor and host from a single pretreatment blood sample that can be utilized as a decision-making tool when considering options for first-line mNSCLC treatment selection. This non-invasive simple test could be readily incorporated into clinic workflow, offering more personalized information for therapeutic decision-making in first-line treatment of mNSCLC.

Limitations of this study encompass the small sample size and inherent potential biases associated with utilizing real-world data, including possible treatment selection biases exhibited by physicians based on practice patterns, clinicopathological factors, or other unmeasured variables. Tumor mutational burden (TMB) measurement was conducted exclusively using tissue-based methods; however, the specific type of next generation sequencing was at the physicians’ discretion and reported by the participating sites in our observational cohort. Lastly, although both biomarkers have been evaluated extensively in both squamous and non-squamous populations treated with both PD-(L)1 inhibitors and CTLA-4 inhibitors across different lines of therapy, our study was restricted to first-line PD-(L)1 inhibitor-based therapy only and underpowered to compare squamous and non-squamous populations independently.

## Data Availability

All data produced in the present study are available upon reasonable request to the authors and included in the supplemental materials

