## Supplemental Data for "Plasma proteome-based test (PROphetNSCLC) predicts response to immune checkpoint inhibitors (ICI) independent of tumor programmed death-ligand 1(PD-L1) expression and tumor mutational burden (TMB)"

**Supplemental Figure 1.** TMB and PROphetNSCLC correlation with outcomes in both PD-(L)1 monotherapy and PD-(L)1 inhibitor combined with chemotherapy

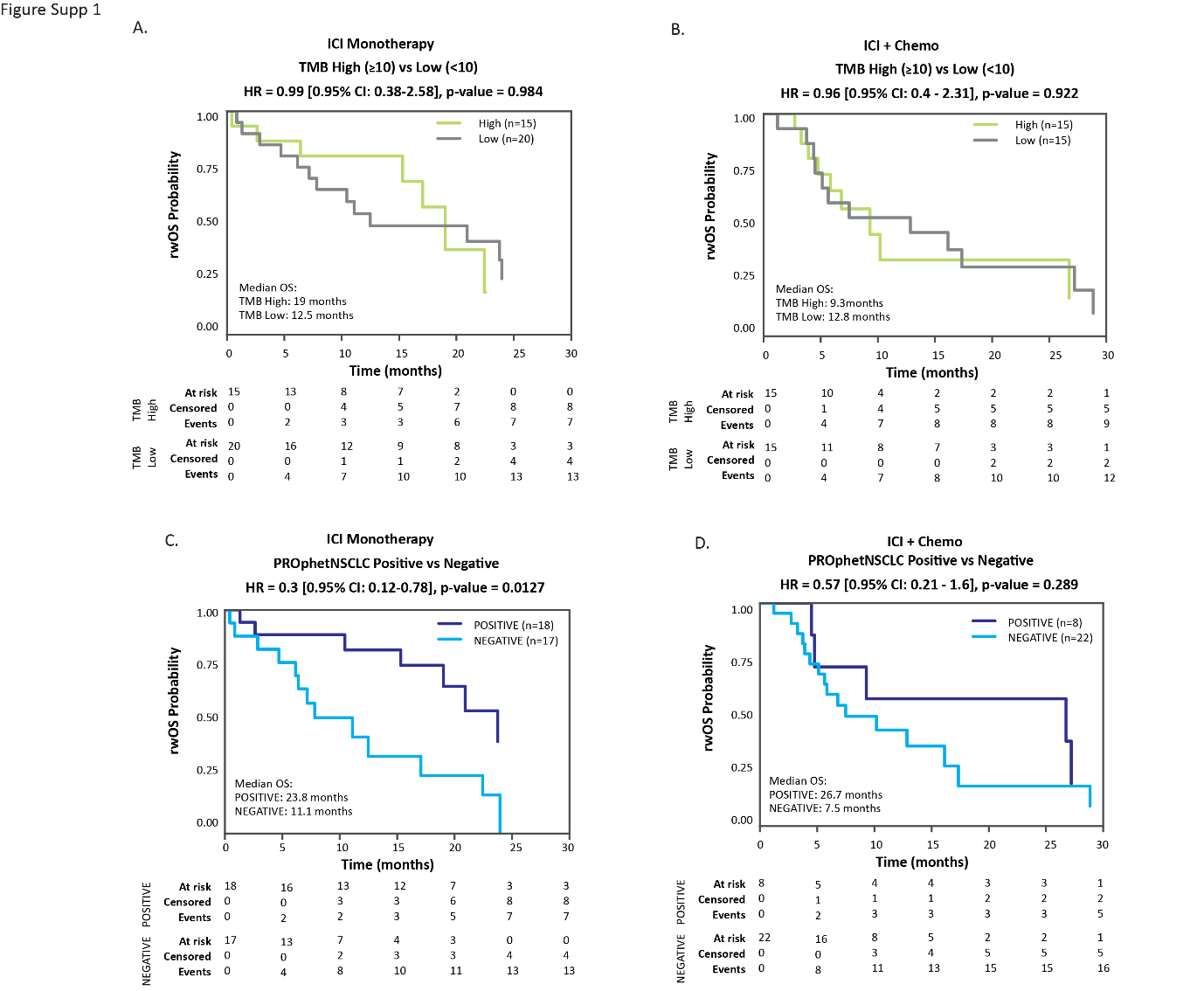

Supplemental Figure 1: This figure illustrates a Kaplan-Meier plot comparing TMB (A+B) and PROphetNSCLC (C+D) as a biomarker in both PD-(L)1 monotherapy treated patients (A+C) versus PD-(L)1 inhibitors in combination with chemotherapy (B+D) given prior data suggesting superior performance of TMB in immunotherapy only treatment regimens.

**Supplemental Table 1.** Characteristics of mNSCLC patients treated with 1L PD-(L)1 inhibitor-based therapy

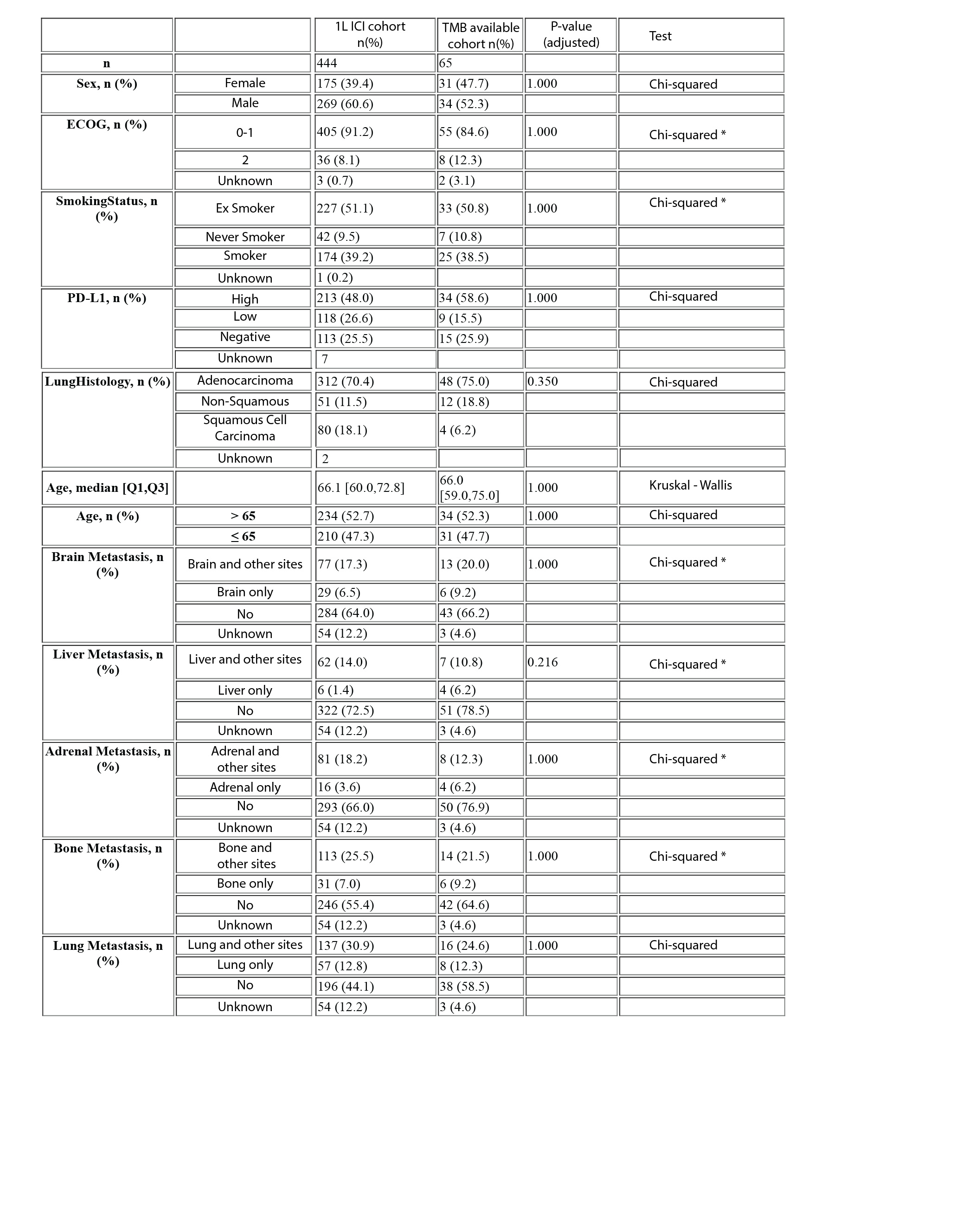

*Chi-square calculations are limited based on small numbers

**Supplemental Table 1. Study cohort:**

[https://github.com/OncoHostPlatform/TMB_analysis_2024/blob/main/Study_cohort_patient_data_with_TMB.csv](https://eur04.safelinks.protection.outlook.com/?url=https%3A%2F%2Fgithub.com%2FOncoHostPlatform%2FTMB_analysis_2024%2Fblob%2Fmain%2FStudy_cohort_patient_data_with_TMB.csv&data=05%7C02%7CYehudab%40Oncohost.com%7C9289000152bf4d91b35b08dccd130471%7C9a5636d7509f4d2184dac95f0112fa85%7C0%7C0%7C638610728759544339%7CUnknown%7CTWFpbGZsb3d8eyJWIjoiMC4wLjAwMDAiLCJQIjoiV2luMzIiLCJBTiI6Ik1haWwiLCJXVCI6Mn0%3D%7C0%7C%7C%7C&sdata=K6kn5aVW1ne6fJ0wrttrVDKh3pUi%2FRar8GJJSE6wjN4%3D&reserved=0)

This table summarizes the dataset the patients with TMB data (n=65) from the Christopoulos et al. study (Christopoulos, Harel, et al., 2024) utilized in this analysis. This table integrates clinical information, model-derived outcomes, tumor PD-L1 and TMB information. The column labeled 'Set' categorizes patients into either the Development Set ('Dev') or the Validation Set ('Val'). Three patients were not included in the Dev or Val set since they couldn’t determine the CB \ NCB status. A separate column, 'Clinical Utility', identifies the patients who underwent in the previous work of the Christopoulos et al. where assessing the clinical implications and were restricted to those who had the PD-L1 score (58 out of 65). The 'TreatmentCombo' column is derived from the 'TreatmentDrugs' column and groups patients based on their treatment regimen: ‘ICI,’ ‘ICI+chemo’ of the patient received ‘chemo’ alone. 'Clinical Benefit' is defined as the absence of disease progression within 12 months post-treatment (for detailed methodology, refer to the paper's Methods section). All patients were treated as first line.
